# The Effect of GDP and Distance on Timing of COVID-19 Spread in Chinese Provinces in 2020

**DOI:** 10.1101/2020.07.19.20157354

**Authors:** Alice Kuan, Mingxin Chen, David Bishai

## Abstract

The geographical spread of COVID-19 across China’s provinces provides the opportunity for retrospective analysis on contributors to the timing of the spread. Highly contagious diseases need to be seeded into populations and we hypothesized that greater distance from the epicenter in Wuhan, as well as higher province-level GDP per capita, would delay the time until a province detected COVID-19 cases. To test this hypothesis, we used province-level socioeconomic data such as GDP per capita and percentage of the population aged over 65, distance from the Wuhan epicenter, and health systems capacity in a Cox proportional hazards analysis of the determinants of each province’s time until epidemic start. The start was defined by the number of days it took for each province to reach thresholds of 3, 5, 10, or 100 cases. We controlled for the number of hospital beds and physicians as these could influence the speed of case detection. Surprisingly, none of the explanatory variables had a statistically significant effect on the time it took for each province to get its first cases; the timing of COVID-19 spread appears to have been random with respect to distance, GDP, demography, and the strength of the health system. Looking to other factors, such as travel, policy, and lockdown measures, could provide additional insights on realizing most critical factors in the timing of spread.

## Introduction

The global COVID-19 pandemic has transformed the world economically and socially. The original epicenter in Wuhan, China slowly lifted lockdown regulations and allowed its citizens to return to normal life in March. Since it was the first country to face the COVID-19 pandemic, China may hold clues to social and geographical factors that affected the spatial spread of the epidemic. This will have implications for other countries now concerned with limiting cross-border introductions of new cases. Recent literature on the spread of the pandemic has highlighted the crucial role of the capacity of health systems – specifically, a sufficient health labor force and adequate volume of intensive care unit (ICU) hospital beds. Italy was one of the earlier well-known examples where severe health worker and ICU bed shortages led to a spike in cases and deaths (Armocida et al., 2020). In Spain, underinvestment leading to insufficient numbers of health professionals and ICU beds contributed to the country’s struggled COVID19 response (Legido-Quigley, Mateos-García, et al., 2020). Lack of bed capacity was also cited as important lessons from Singapore, Hong Kong, and Japan (Legido-Quigley, Asgari, et al., 2020). In addition, past research on other infectious disease outbreaks have observed significant relationships between population density and disease incidence, as was the case of hand, foot, and mouth disease (HFMD) in China (Hu et al., 2012).

Other articles have also examined the interaction between macroeconomic factors and the spread of infectious diseases. The prevalence of tuberculosis in EU member states in 2010 had an inverse relationship with their GDP levels, accounting for wealth distribution within each country (Semenza et al., 2010). More generally, macroeconomic factors such as national distribution of wealth or GDP per capita, affects the quality of population health (Subramanian et al., 2002). Poor macro socio-economic conditions provide more opportunities and environments in which infectious diseases spread. Economic growth can also alter the prevalence of infectious agents and lead to aging populations, which heightens vulnerability to disease (Ceddia et al., 2013).

Literature about COVID-19 spread in China has found significant correlations between transportation links between Wuhan and other cities and COVID-19 caseloads. One study applied Pearson’s correlation analysis and found daily frequencies of flights, bus trips and train trips was correlated to cumulative COVID-19 cases (Zheng et al., 2020). Another article found an inverse square relationship between other regions distance from Wuhan and their cumulative number of cases, with a correlation coefficient of 0.267 for places in China and -0.197 for the rest of the world (Biswas & Sen, 2020). International studies show that proximity to China seems to have affected the number of days from the first reported cases in China until COVID-19 arrives in another country (Adegboye et al., 2020).

These articles provide helpful insights into associations between several factors and the volume of cases. They also reveal, however, additional opportunities to control for other variables – including macroeconomics or health systems capacity as highlighted above. The volume of health workers or the number of elders within a population could influence the speed of spread, thus serving as confounders. In addition, past studies did not address the timing of each location’s cases. We aim to include these considerations in our paper and re-examine the question of geospatial spread of COVID-19 across provincial borders in China. We apply event history analysis to determine the effects of province-level GDP per capita and distance from Wuhan on the time it took for each Chinese province to reach its first wave of cases, measured as elapsed number of days between January 22, 2020 and the occurrence of various thresholds of 3, 5, 10, or 100 cumulative cases. We control for health systems capacity and population factors, including the number of doctors and hospital beds per 1000 people in each province, proportion of the population aged over 65, and other variables that are delineated below.

## Methods

The econometric model included 10 explanatory variables described in Table 1. Data on the number of cases by province were drawn on May 26, 2020 from a GitHub repository managed by the Johns Hopkins Center for Systems Science and Engineering. Data on provincial economic and demographic variables are in the public domain.

**Table 1:**
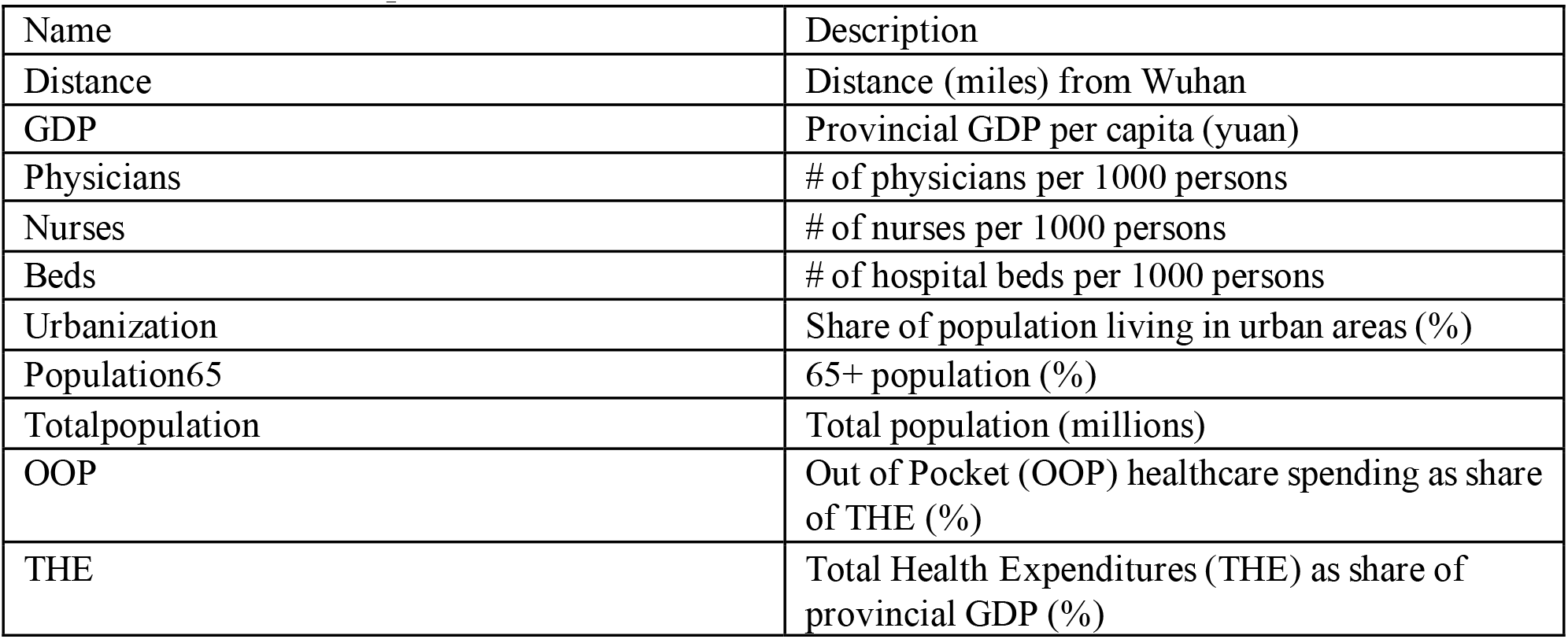
Variable Descriptions.

Our analysis used the Cox proportional hazards model shown below:

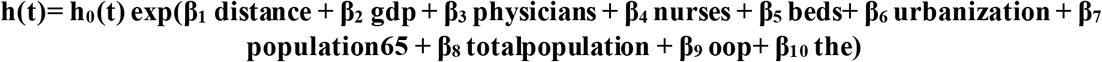

where h(t) is the hazard of reaching a COVID-19 threshold between time 0 (January 22) and time t, and h_0_ is a common baseline hazard which is modified by the covariates listed below.

Table 3 in the Appendix provides a correlation matrix that gives more insight into the general relationship between each independent variable at the province level. The matrix shows that higher GDP per capita is associated with a larger proportion of elders, consistent with literature discussing how economic growth could lead to aging populations. Higher GDP is also associated with more healthcare workers, while it correlates with less OOP and THE. This could be because more wealth per capita leads to a healthier population, lessening the need for health spending. Furthermore, a wealthier region could have more economic opportunity, such as in education which allows people to become doctors or nurses.

**Table 2:** Results of Cox Regressions using Different Thresholds as Dependent Variables.

|  | Alternative Definitions of Failure Time Based on Cumulative Cases |  |  |  |
| --- | --- | --- | --- | --- |
| Independent Variables | Time until 3 Cases | Time until 5 Cases | Time until 10 Cases | Time until 100 Cases |
| distance | -0.000262<br>(-0.620) | -0.000297<br>(-0.664) | 0.000131<br>(0.502) | 0.000100<br>(0.392) |
| GDP | 6.96e-06<br>(0.182) | 7.79e-06<br>(0.232) | 5.00e-06<br>(0.161) | -1.79e-05<br>(-0.571) |
| physicians | -0.286<br>(-0.272) | -0.335<br>(-0.348) | -0.815<br>(-0.894) | -0.120<br>(-0.150) |
| nurses | 0.0461<br>(0.0590) | -0.123<br>(-0.169) | 0.122<br>(0.177) | 0.338<br>(0.458) |
| beds | 0.00422<br>(0.00893) | -0.0587<br>(-0.132) | 0.000304<br>(0.000746) | -0.317<br>(-0.724) |
| urbanization | -0.0612<br>(-0.704) | -0.0402<br>(-0.512) | -0.0111<br>(-0.160) | 0.0284<br>(0.437) |
| population65 | -0.0385<br>(-0.214) | -0.0603<br>(-0.366) | 0.0835<br>(0.566) | 0.134<br>(0.949) |
| totalpopulation | -0.0103<br>(-0.673) | -0.0120<br>(-0.884) | -0.0100<br>(-0.805) | 0.000213<br>(0.0216) |
| OOP | 0.104<br>(1.008) | 0.105<br>(1.122) | 0.0455<br>(0.546) | -0.0128<br>(-0.141) |
| THE | -0.0645 | -0.0497 | -0.0137 | -0.115 |
|  | (-0.230) | (-0.191) | (-0.0595) | (-0.433) |
| Observations | 3,844 | 3,844 | 3,844 | 3,844 |
| Number of Provinces<sup>1</sup> | 9 | 14 | 26 | 30 |
| F | 4.878 | 5.211 | 3.038 | 3.127 |
| P | 0.899 | 0.877 | 0.981 | 0.978 |

z-statistics in parentheses \*\*\* p<0.01, \*\* p<0.05, \* p<0.1

**Table 3:** Correlation Matrix of Independent Variables.

|  | distance | gdp | physicians | nurses | beds | urbanization | population65 | totalpopulation | oop | the |
| --- | --- | --- | --- | --- | --- | --- | --- | --- | --- | --- |
| distance | 1 |  |  |  |  |  |  |  |  |  |
| gdp | -0.23493 | 1 |  |  |  |  |  |  |  |  |
| physicians | -0.05184 | 0.682382 | 1 |  |  |  |  |  |  |  |
| nurses | -0.19317 | 0.601583 | 0.746449 | 1 |  |  |  |  |  |  |
| beds | 0.15041 | -0.28409 | 0.066605 | 0.197905 | 1 |  |  |  |  |  |
| urbanization | -0.28485 | 0.885996 | 0.624364 | 0.636315 | -0.17849 | 1 |  |  |  |  |
| population65 | -0.24067 | 0.286163 | 0.230847 | 0.272338 | 0.400414 | 0.406788 | 1 |  |  |  |
| totalpopulation | 0.083554 | 0.024233 | -0.12189 | -0.04802 | 0.059953 | 0.016893 | 0.432973 | 1 |  |  |
| oop | 0.014729 | -0.25408 | -0.16768 | -0.05936 | 0.36338 | 0.095631 | 0.440329 | 0.353388 | 1 |  |
| the | 0.222221 | -0.51529 | -0.12223 | -0.14877 | 0.243369 | -0.54769 | -0.47581 | -0.54549 | -0.37311 | 1 |

We used Stata IC16 to estimate the proportional hazards model. For sensitivity analysis we tested four alternative definitions of the failure time. In each case, we started observations on January 22 and declared failure (i.e. epidemic onset) when a province reached its first 3, 5, 10, or 100 cases. Each independent variable coefficient indicates the effect of the variable on the amount of time a province takes to reach a threshold. Several provinces reported more than 3 or 5 cases on January 22 and could not contribute meaningful observations to the event history analysis with low thresholds. Therefore, repeating the model with higher thresholds such as 100 cases ensured that the results would account for a wider number of provinces.

## Results

From Table 2, we see that the coefficients on provincial GDP and distance have no statistically significant effect on the number of days it takes for a province to reach any of the thresholds. This disproves our initial hypotheses that distance and macroeconomic factors would alter the speed of COVID-19 spread. We find that similarly and remarkably, none of the other variables significantly affect the time until the pandemic starts in a province. The F-statistic shows that the joint significance of all explanatory variables together is compatible with the null hypothesis that nothing explains the time to failure.

Figures 1a-d provide Kaplan-Meier curves displaying the probability of survival – that is, cases staying below the threshold – accounting for all provinces over time. Each step represents a failure event, or at least one province reaching or surpassing the threshold. The greater the number of days from January 22, the greater the number of cases and therefore the lower survival probability. We see from Figures 1a-b that the probability decreases rapidly for thresholds of 3 and 5 cases, likely because they are very low minimums to surpass. On the other hand, the probability decreases at a slower pace for thresholds of 10 and 100, particularly for the latter because it took longer for some provinces than others to surpass 100 cases. Figure 1d reflects this by showing more variation across provinces in the timing of events.

**Figure 1:**
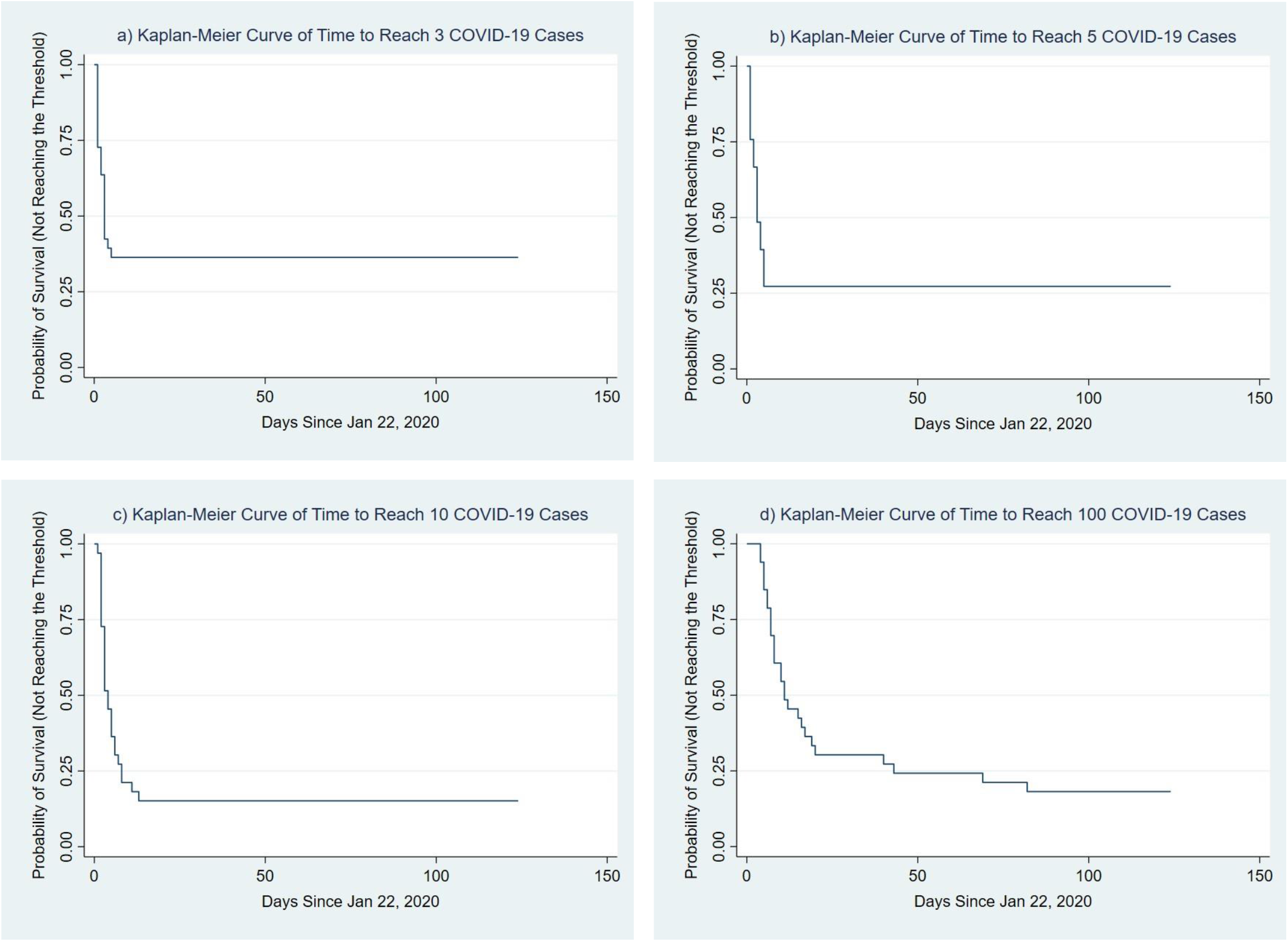
Kaplan-Meier Curves of Time to Reach Four Different Thresholds.

## Discussion

Our results are inconsistent with recent literature analyzing the effects of distance on the severity of the COVID-19 epidemic, as well as with older studies showing that macroeconomic and socioeconomic variables play a role in disease spread. We intend to further research in these areas and provide suggestions for future study.

There are a few reasons that our results could differ. Unlike recent studies, our model controls for population variables and socioeconomic factors when assessing distance from Wuhan, as we include GDP, population, and health worker data in our model. In addition, our methodology takes advantage of provincial differences in the timing of each province-level epidemic, rather than examining the magnitude of the count of cases on any given date. This allows our research to bear a unique dependent variable, providing analyses specific to the context of the number of days until epidemic start which, to our knowledge, was not the primary focus of other studies.

Another reason could be that the Chinese New Year festival diaspora on January 31, 2020 was able to seed every province with a sufficient number of infected travelers at approximately the same time. An estimated 5 million people left Wuhan before the start of the travel ban on January 23, and a third of these travelers moved to locations outside Hubei province (Chen et al., 2020). Our results would suggest that these roughly 1.6 million outbound travelers included enough latently infected individuals to seed other provinces with COVID-19. For this scenario to be compatible with our findings, distance from Wuhan would not serve as a factor that would delay the arrival of the New Year’s celebrants, and neither GDP nor health systems capacity had bearing on the containment of the illness. Thus, the provinces appear to have sufficiently equitable economic states so that provincial GDP is an immaterial factor in the timing and initial spread of the epidemic. This could be a finding unique to the country, but there are other considerations as well, such as the timing of the lockdown measures and subsequent changes in crossprovince travel that could serve as other research areas. Future studies on in-country travel data and lockdown policies, while controlling for the variables presented in this paper and other relevant socioeconomic factors, may provide a new angle of observing the COVID-19 spread within different countries.

## Data Availability

The data on cumulative COVID-19 cases in Chinese provinces is publicly available from the JHU CSSE Github repository. Data on province-level variables is available from Beijing University.

https://github.com/CSSEGISandData/COVID-19

## Acknowledgements

We would like to acknowledge help from PeiPei Chang at Beijing University for gathering province level data, and Katelyn Jisoo Yoon from the World Bank for helpful comments.

## Footnotes

1 Number of Provinces indicates how many provinces were included in the analysis for each threshold; in other words, how many provinces had not yet reached the threshold on Jan 22 ^nd^, 2020. There are 31 provinces in total. The number of provinces that had already reached or surpassed the threshold by that day are thus 22, 17, 5, and 1 respectively in order of the columns from left to right.

